# Comparison of Time- and Frequency-Domain Methods for Assessing Brain Compliance in Brain-Injured Patients Monitored With Intraparenchymal Intracranial Pressure Sensors

**DOI:** 10.64898/2025.12.10.25341990

**Authors:** Ghibaudo Valentin, Elhadjene Nory, Percevault Gwendan, Bapteste Lionel, Gobert Florent, Carrillon Romain, Bodonian Carole, Contard Florian, Mayras Anaïs, Ravachol Alexandre, Manet Romain, Dailler Frédéric, Rithzenthaler Thomas, Garcia Samuel, Balança Baptiste

## Abstract

**Introduction:** Intracranial pressure (ICP) monitoring is commonly used in neuro-intensive care, but its utility may be limited by a suboptimal use. The brain pressure-volume relationship, a potential predictor of neurological health, is now approached using time-domain methods, which can be challenging to implement. Frequency-domain methods may offer an alternative, but their relationship with time-domain metrics remains unclear. This study compares time- and frequency-domain methods for assessing brain compliance and evaluates their real-time usability.

**Methodology:** A monocentric, prospective observational study was conducted in the neurological ICU of the Hospices Civils de Lyon, France, to evaluate markers of brain compliance. Adult patients with brain lesions requiring multimodal monitoring were included. Continuous high-density physiological data, including ICP, arterial blood pressure (ABP), end-tidal CO₂, and electrocardiogram (ECG), were collected for analysis. Some spontaneous ICP rise events were automatically detected based on heuristic criteria and used as brain compliance challenges to compare the co-evolution of metrics across multiple time windows. Time-domain (pulse shape-related metrics) and frequency-domain analyses (examining heart and respiratory components in ICP) were computed to assess the intracranial pressure-volume state. Statistical analyses were performed using linear mixed-effects modeling, adjusting for vasoactive and sedative medications and Spearman correlations.

**Results:** The study included 66 patients, with a mean age of 49.16 [38.38, 57.58] years. A total of 518 spontaneous ICP rise events were detected in 56 patients. Our findings revealed that: 1) frequency-domain metrics strongly correlated with time-domain metrics during these challenges (r > 0.8, p < 0.001), 2) frequency-domain metrics were significantly drastically less computationally demanding, and 3) the impact of the heart on ICP showed a significant correlation with the P2/P1 ratio (r = 0.391, p < 0.001) and other potential markers of brain compliance. In contrast, the impact of respiration on ICP was only marginally correlated with these markers.

**Conclusions:** Frequency-domain analyses exploring the impact of cardiac activity on ICP map provide similarly informative value to more complex machine learning-based tools, but with the advantage of being much less computationally demanding. This makes this approach particularly suitable and intuitive for real-time clinical monitoring in hospital settings, where computational resources are often limited.

**Authors statement:** Initials: VG, NE, GP, LB, FG, RC, CB, FC, AM, AR, RM, FD, TR, SG, BB

- Conception and Design: VG, NE, BB
- Data Collection : VG, NE, LB, GP, TR, FG, RC, CB, FC, FD, BB
- Data Analysis and Interpretation: VG, BB
- Methodology Development: VG, BB, SG
- Manuscript Drafting and Writing: VG, BB
- Supervision and Project Oversight: BB
- Funding Acquisition: BB, VG
- Visualization: VG, BB
- Critical Review and Editing: NE, GP, LB, FG, RC, CB, FC, AM, AR, RM, FD, TR, SG, BB
- Approval of the Final Version: All
- Software Development: VG, SG
- Ethical Approval and Regulatory Compliance: BB

## 1. Introduction

Severe acute brain injuries encompass a heterogeneous group of conditions, including traumatic brain injury (TBI) or aneurysmal subarachnoid hemorrhage (aSAH), which necessitate admission to a neuro-intensive care unit (neuro-ICU). The optimization of vital functions and the body’s homeostasis, as well as the early detection of new insults, are key components of critical care management, in addition to treating the underlying precipitating condition. Thus, cerebral metabolism and cerebral blood flow can be optimized, notably searching for the optimal cerebral perfusion using monitoring data derived from measurements such as intracranial pressure (ICP), brain tissue oxygen pressure (PbtO2), and microdialysis (Helbok et al., 2017; Okonkwo et al., 2017; Tas et al., 2021). The burden of high ICP or inappropriate cerebral perfusion pressure (CPP) is associated with a poorer prognosis (Dias et al., 2015; Güiza et al., 2015; Tas et al., 2021; Vik et al., 2008). However, the usefulness of ICP monitoring and the optimal CPP and ICP targets remain controversial. Therefore this monitoring is not undertaken in all brain-injured patients, while ICP monitoring seems to improve mortality and change the therapeutic intensity level (R. M. Chesnut et al., 2012; Robba et al., 2021).

One potential way to enhance the clinical value of ICP monitoring is by deriving brain compliance, which reflects the intracranial tissue’s ability to accommodate volume changes. According to the Langfitt curve, high brain compliance is characterized by minimal ICP changes in response to intracranial volume variations (Brasil et al., 2021; Godoy et al., 2023). Therefore, intracranial compliance may serve as a more reliable clinical target than ICP alone (Godoy et al., 2023). However, brain compliance cannot be directly measured at the bedside and must instead be approximated through various ICP signal features. Several metrics have been proposed to assess brain compliance, most of which are based on ICP waveform analysis (Brasil et al., 2021; Czosnyka et al., 1988; Kazimierska et al., 2023). Since some of the intracranial compartments (e.g., cerebrospinal fluid, arterial, and venous compartments) naturally undergo volume variations due to respiratory and cardiac cycles (Dreha-Kulaczewski et al., 2015; Söderström et al., 2025), ICP changes induced by these physiological rhythms have also been used to approximate such a pressure-volume relationship (Spiegelberg et al., 2020).

To date, many proposed compliance metrics have been developed using time-domain analyses (i.e., examining how a signal changes over time, while frequency domain analysis focuses on its frequency components). However, these approaches often rely on numerous parameters or machine learning models (Legé et al., 2023; Mataczyński et al., 2022), which can be computationally demanding and difficult to implement in a real-time neuro-ICU environment. Frequency-domain analysis presents a potential alternative that could provide a computationally efficient means of extracting compliance-related information and offer deeper insights into ICP dynamics by examining multiple frequency components simultaneously.

No research has yet comprehensively examined the statistical relationships between time-domain and frequency-domain techniques used to evaluate brain compliance. The ideal marker at the bedside should require low computational resources and undergo significant changes in response to cerebral volume fluctuations. Comparing time and frequency domain analyses with these two aspects could help determine the most appropriate ones. In the neuro-ICU, patients naturally experience spontaneous intracranial compliance challenges, often manifesting as transient ICP elevations due to various factors, including patient mobilization, metabolic changes, therapeutic interventions, and spontaneous slow-waves (Kasprowicz et al., 2016). In this study, we aim to compare the co-evolution of multiple time- and frequency-domain methods during these spontaneous brain ICP challenge events and evaluate their practical computational usability.

## 2. Materials and methods

### Study Design

We conducted a monocentric, prospective observational trial in the neurological intensive care unit (ICU) of the Hospices Civils de Lyon, France. The MultiICU study was approved by the *Comité scientifique et éthique des Hospices Civils de Lyon* (ethical committee IRB 0013204, n°713, on the 06/07/2023) and declared to the French national information and liberty commission (CNIL No. 25_5713). For this observational analysis, patients or their relatives were informed of their participation in this investigation, and their written consent was not considered as necessary by our local ethical committee. This study investigates several aspects of multimodal neurological monitoring after acute brain injuries, such as aneurysmal subarachnoid hemorrhage (SAH), severe traumatic brain injury (TBI) or intraparenchymal hematoma. We retrospectively and prospectively collected data from digital medical records since January 2020.

### Study population

This study included adult men and women (≥18 years of age) admitted to the neurological ICU with poor grade SAH, severe TBI, or other severe brain conditions, defined by an initial Glasgow Coma Scale score of less than 8. Eligibility required participants to undergo multimodal monitoring—such as expired CO2 monitoring, invasive arterial blood pressure (ABP) measurement, and intraparenchymal intracranial pressure (ICP) monitoring (through a bold or a surgical interventions, including hematoma evacuation, aneurysmal clipping, or external ventricular drainage). Exclusion criteria for this analysis included patients without available CO2 monitoring data due to technical problems.

### Data acquisition

High-resolution monitoring data were collected using the Moberg CNS Monitor (CNS-320, MICROMED-NATUS, USA). Intracranial pressure (ICP) was measured using an intraparenchymal probe (Pressio, Sophysa, Orsay, France) inserted into the frontal cortex. Arterial blood pressure (ABP) was measured via a radial or femoral line using standard monitoring kits (ICUMedical, California, USA). Respiratory data were obtained by measuring the partial pressure of CO2 in the endotracheal tube (Infinity® MCable™ Mainstream CO2, Dräger Médical, Antony, France). Electrocardiographic (ECG) data were recorded using a thoracic 5-lead system. All signals were continuously recorded and displayed by the IntelliVue MX750 monitor (Phillips, SURESNES, France) connected to the CNS monitor with the following sampling rates: 125 Hz for ICP and ABP, 500 Hz for ECG, and between 60 Hz and 120 Hz for CO2. Continuous clinical and therapeutic data were also collected through extraction from the IntelliSpace Critical Care and Anesthesia (ICCA, Phillips, SURESNES, France) software, which is used in the unit for prescription management.

### Data processing

Data processing was done using Python language. Codes are available in the link provided in the code availability statement at the end of the manuscript. Several steps were taken from the raw data to statistical analysis as follows:

#### ○ From raw files to python: pycns

We developed a custom Python toolbox called *pycns* (available at https://github.com/samuelgarcia/pycns specifically designed to convert raw Moberg CNS Monitor files into a Python-compatible format. This toolbox allowed us to generate continuous time series data for CO2, ICP, ABP, and ECG levels for each patient in a NumPy format (Harris et al., 2020), facilitating scientific computations and analysis.

#### ○ Slow ICP rises detection and dividing into multiple windows

The purpose of this study is to compare time- and frequency-domain methods in their approach to assessing brain compliance. Specifically, we aim to evaluate and contrast these methods during spontaneous brain compliance challenges. To achieve this, we developed an automated detection system for identifying spontaneous increases in intracranial pressure (ICP), followed by an automated computation of compliance metrics. A graphical overview of the process is proposed in the Figure *1*.

**Figure 1:**
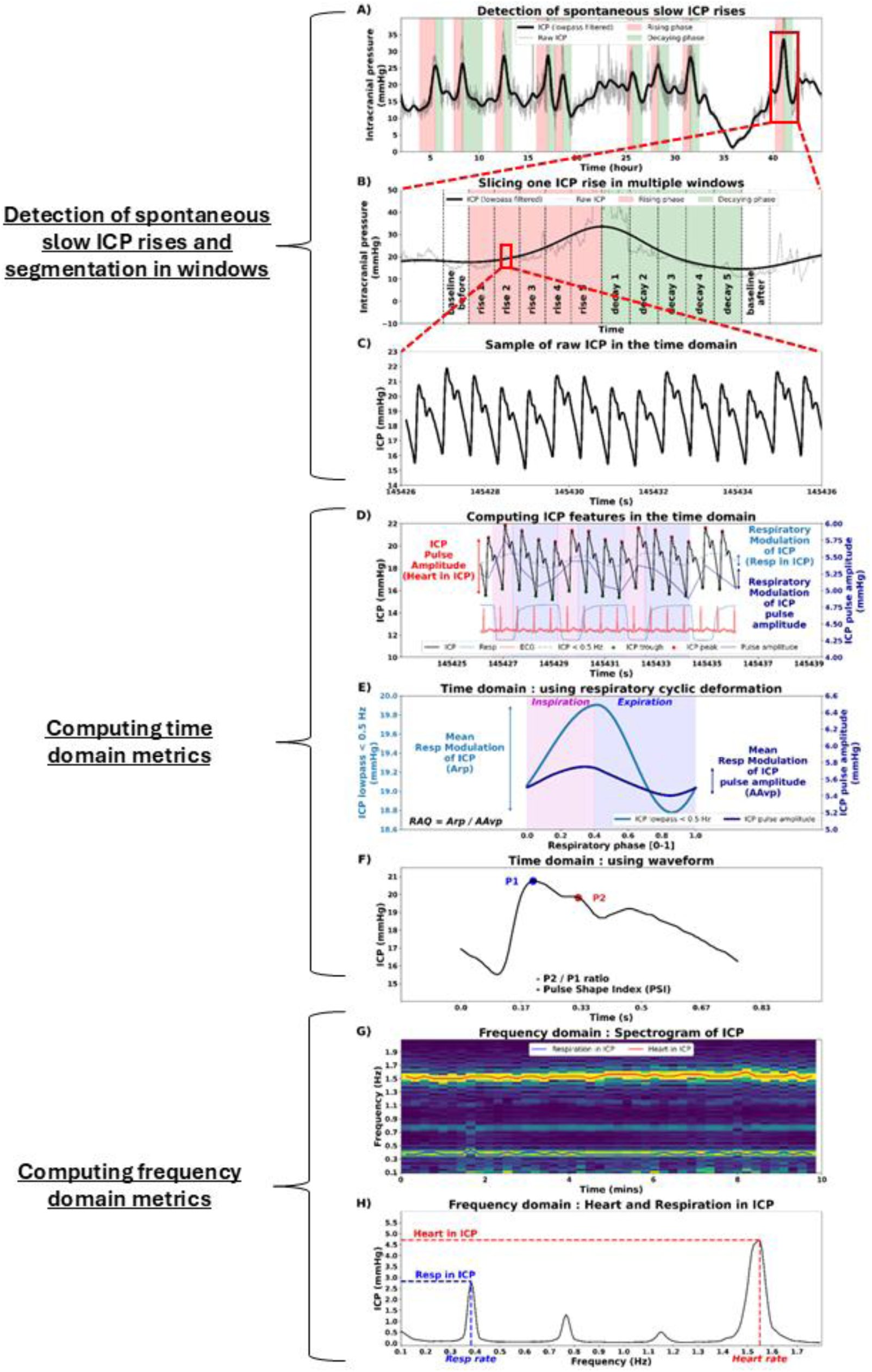
Graphical overview of metrics computation. A) Example of detections of spontaneous slow ICP rise events. On this sample of 48 hours of ICP signal, 9 events following the detection criteria were detected on a smoothed ICP trace (black line). Their rising phase (period while ICP increases) is displayed in red, while their decaying phase (period while ICP decreases) is displayed in green. B) Example of a time course of ICP during one slow ICP rise. The period corresponds to the last event displayed in panel A (red box). Each event is sliced in 12 windows, during which compliance metrics are computed. C) A small sample of ICP is displayed in the time domain (red box in panel B). D) Some metrics related to brain compliance are computed directly in the time domain. ICP pulse amplitude (red arrow) measures how the heart impacts the ICP (see the corresponding cardiac activity on ECG trace). Respiration can impact both the ICP level itself (light blue arrow showing the modulation by respiration of an ICP smoothed signal < 0.5 Hz) and the ICP pulse amplitude (dark blue arrow showing the modulation by respiration of the ICP pulse amplitude trace). Such phase-amplitude coupling is evidenced by the displaying of inspiration and expiration phases in green and red vertical spans that we were able to detect on the endotracheal CO2 levels (light blue continuous trace). E) To precisely characterize the impact of respiration on both ICP level itself and ICP pulse amplitude, each of their respiratory epoch time basis was rescaled to a respiratory phase basis, allowing for an inter-epoch average (see Ghibaudo et al., 2023; Roux et al., 2006). The obtained averaged respiratory modulation of ICP by respiration corresponds to a more accurate extraction of the A_rp_ component defined previously (Spiegelberg et al., 2020) while the averaged respiratory modulation of the ICP pulse amplitude corresponds to a more accurate extraction of the AA_vp_ component defined in the same article. Therefore, we computed a more accurate RAQ (A_rp_ / AA_vp_). F) Time-domain metrics based on pulse waveforms were also computed : P2/P1 ratio and Pulse Shape Index using already trained machine-learning models (Legé et al., 2023; Mataczyński et al., 2022). G) The impact of respiration and heart on ICP was also computed in the frequency-domain by tracking the amplitude of the respiratory (blue) and heart (red) components on a spectrogram of ICP. H) Averaging the spectrogram displayed in G along the time axis allows for a better representation of the amplitude of the heart and respiratory components in ICP.

The detection of spontaneous increases in ICP was done using the following criteria:

1. The rising phase must last for at least 15 minutes.
2. The ICP must increase by at least 5 mmHg.
3. The ICP must exceed a threshold of 20 mmHg.
4. The start of the rising phase must not exceed 20 mmHg.

We applied these detection criteria to the first derivative of a filtered version of the raw ICP signal. To obtain the filtered ICP signal, we used a 10th-order Bessel low-pass filter with a cutoff frequency of 0.00111 Hz, corresponding to a 30-minute period duration. An example of the detected events is presented in Figure *1*A.

The rising and decaying phases of each ICP event were divided into five equal-duration windows (see Figure *1*B), allowing us to compute average values for each compliance metric within each window. For statistical analysis, we compared the compliance values obtained in each window to baseline values. Two baseline windows were defined for this purpose :

- A pre-rising phase baseline window of equal duration to the rising-phase windows.
- A post-decaying phase baseline window of equal duration to the decaying-phase windows.

These baseline windows provided reference points for evaluating changes in compliance during the events.

#### ○ Computing time domain metrics

Sixteen metrics were initially computed directly in the time-domain (see Figure *1* C, D, E, F). The timestamps of detected slow ICP rises were then utilized to extract a central tendency measure (mean or median) for each window / event / patient. Some of these metrics are closely associated with compliance assessments, while others are more relevant to evaluating cerebral autoregulation.

1. Median ICP (mmHg): The median values of the ICP-filtered signal corresponding to each window were calculated.
2. ICP Pulse Amplitude (mmHg): ICP pulses induced by cardiac activity were identified in a filtered version of the raw ICP time series using a 4th-order Butterworth band-pass filter with cutoff frequencies of 0.1 to 10 Hz. The detection was performed with the *signal. findpeaks()* function taken from the *SciPy* toolbox, enabling the extraction of the raw ICP values at the trough preceding each pulse and at the peak of each pulse (Virtanen et al., 2020). The difference between these peaks and troughs values represents the pulse amplitude in mmHg (see Figure *1*D). For each window, a single pulse amplitude value was obtained by calculating the median of the amplitudes of all pulses within the current window.
3. Respiratory Modulation of ICP Pulse Amplitude: This metric measures the impact of the respiratory phase on the amplitude of the cardiac induced pulse in ICP. It can be compared to the AA_vp_ component of the RAQ index described in Spiegelberg et al., 2020 (Spiegelberg et al., 2020). Our method should provide a more accurate measurement of respiratory-induced changes in the ICP signal, as it leverages the respiratory signal (CO2 in the endotracheal tube) to precisely determine the timing of each respiratory cycle. Indeed, we first detected all the respiratory cycles from the endotracheal CO2 trace. Then, we interpolated the full ICP Pulse Amplitude time series in order for that to be equally sampled as the CO2 trace. We then used the *deform_traces_to_cycle_template()* of the *physio* toolbox (Ghibaudo et al., 2023) to proceed to a cyclical deformation of the ICP Pulse Amplitude resampled time series signal based on respiratory cycle timestamps. This process provided an ICP Pulse Amplitude time series for each respiratory cycle (see Figure *1* D, E). From this, the peak-to-peak range of each epoch was extracted to get one value of modulation by the respiratory cycle. For each window, a single value was computed as the median of all ICP Pulse Amplitude Respiratory-Induced Variations identified within that window (see Figure *1*E).
4. Respiratory Modulation of ICP: This metric measures the impact of the respiratory phase on the amplitude of the ICP. It can be compared to the A_rp_ component of the RAQ index described in Spiegelberg et al., 2020 (Spiegelberg et al., 2020). The same process of respiratory cyclical deformation described previously is applied but directly on a filtered ICP trace (4^th^ order Butterworth lowpass filter with a cutoff frequency of 0.5 Hz to remove components faster than respiration). The peak-to-peak range of each epoch was extracted, and the median value of each window was computed according to their timestamps (see Figure *1* D, E).
5. RAQ_2: The ratio Respiratory Modulation of ICP / Respiratory Modulation of ICP Pulse Amplitude was computed for each respiratory cycle, and the median value of each window was kept, providing a second version of the RAQ index described in the literature (Spiegelberg et al., 2020), but possibly more accurate due to the use of respiratory signals (see Figure *1*E).
6. P2P1 ratio: A deep learning-based automated method, previously published (Legé et al., 2023), was utilized to compute a complete time series of P2/P1 ratios. The P2/P1 ratio has been shown to vary in relation to compliance (Brasil et al., 2021). For each window, a single representative ratio was determined by calculating the median of the P2/P1 ratios for all measurable pulses within that window (see Figure *1*F).
7. Pulse Shape Index (PSI): A deep learning-based automated method, previously published (Mataczyński et al., 2022), was utilized to compute a complete time series of Pulse Shape Index (PSI). It has also been shown to be associated with brain compliance (Kazimierska et al., 2023). For each window, a single PSI value was calculated as the mean of all PSI values obtained within that window (see Figure *1*F).
8. Median ABP (mmHg): The median values of the ABP signal corresponding to each window were calculated.
9. Median CPP (mmHg): The median ABP value of each window was subtracted from the corresponding median ICP value to compute the cerebral perfusion pressure (CPP).
10. ABP Pulse Amplitude (mmHg): The same process described in the 2^nd^ paragraph of the current section was applied to the ABP signal but with a 1st-order Bessel band-pass filter with cutoff frequencies of 0.3 to 10 Hz. For each window, a single pulse amplitude value was obtained by calculating the median of the amplitudes of all pulses within the current window.
11. Respiratory Modulation of ABP Pulse Amplitude: The same process described in the paragraph “Respiratory Modulation of ICP Pulse Amplitude” was applied to the ABP signal. The only changes concerned the ABP pulse detection that was done using the same method but on a 1^st^ order Bessel bandpass (0.3 to 10 Hz) ABP filtered signal. This measure has been related to the hemodynamic state of the patient, notably the preload dependency (Bronzwaer et al., 2015), a condition that could interfere with brain pressure-volume interactions.
12. Respiratory Modulation of ABP: The same process described in the paragraph “Respiratory Modulation of ICP” was applied to the ABP signal.
13. RAQ_ABP: The ratio Respiratory Modulation of ABP / Respiratory Modulation of ABP Pulse Amplitude was computed for each respiratory cycle, and the median value of each window was kept.
14. The Pressure-Reactivity Index (PRx): The PRx was calculated as a moving correlation coefficient between 30 consecutive samples of values for ICP and ABP averaged for a period of 10 seconds (overlapping 80% between each moving window) on the available time series of ICP and ABP. For each window, a single value was calculated as the median of all PRx values obtained within that window.
15. Heart rate (bpm): We used the *physio* toolbox to detect ECG R peaks and compute the instantaneous heart rate (60 divided by the interbeat interval in seconds). For each window, a single heart rate value was derived by taking the median of all instantaneous heart rate values within that window.
16. Respiratory rate (cpm): We utilized the previously described respiratory cycle detections to compute the instantaneous respiratory frequency (60 divided by the cycle duration in seconds). For each window, a single respiratory rate value was derived by taking the median of all instantaneous respiratory rate values within that window.

#### ○ Computing frequency domain metrics

Three metrics were computed using the frequency domain of the complete ICP time series for each patient. The timestamps of detected slow ICP rises were then utilized to extract a central tendency measure (median) for each window, event, or patient.

To achieve this, we first used the function *scipy. signal. spectrogram()* from the *SciPy* toolbox on the raw ICP signal to compute a spectrogram (Density Spectral Array) representing the power of the entire ICP signal for a patient. The spectrogram was calculated using Tukey windows of 1-minute duration with 50% overlap and expressed in spectral units (mmHg²). The resulting values were square rooted to convert them back to amplitude units in mmHg.

1. Heart in ICP Spectrum: After applying a mask to the spectrogram matrix to isolate the 0.8–2.5 Hz range (the cardiac spectral band), the maximum amplitude values along the frequency axis were extracted, resulting in a time series representing the amplitude of the cardiac component. The timestamps of the detected windows were then used to segment the time series, and the median value was calculated for each window (see Figure *1* G, H). This process theoretically corresponds to the previously described ICP Pulse Amplitude extraction performed in the time domain, but is here obtained in the frequency domain.
2. Respiration in ICP Spectrum: The same process described in the previous paragraph was applied, but with a mask isolating the 0.12–0.60 Hz range, which corresponds to the respiratory spectral band (see Figure *1* G, H) of patients in Neuro-ICU.
3. Heart / Resp Spectral Ratio: The values obtained as described in the previous two paragraphs were divided to calculate the ratio of Heart Amplitude to Respiration Amplitude.

#### ○ Computational resource requirements

To evaluate the practical usability of the metrics, we implemented a systematic performance measurement framework. We selected five representative metrics—Heart in ICP time domain (ICP Pulse Amplitude), Resp in ICP time domain (Respiratory Modulation of ICP), Heart and Respiration in ICP Spectrum, PSI, and P2P1_ratio—as they could summarize the ones described in the previous paragraph. These functions were tested on one-hour segments of data and iterated over all the available datasets from patients. To measure computational performance, we implemented a profiling routine that captures execution time, CPU usage, and memory consumption. A separate monitoring thread continuously recorded the CPU load and memory footprint of each function while it executed. CPU utilization was sampled at 0.1-second intervals, and memory usage was tracked by monitoring resident set size (RSS). Each function was executed with appropriate input arguments, and the peak CPU usage, maximum memory consumption, and total execution duration were recorded. The results were aggregated into a structured dataset for further analysis.

The experiments were conducted in two different computing environments to evaluate the computational load under varying hardware conditions. The first system was a high-performance workstation equipped with an Intel® Xeon® Silver 4210 CPU (2.20 GHz, 40 logical processors, 10 cores per socket, 2 sockets) with a maximum clock speed of 3.2 GHz. The second system was a lower-powered machine, similar to those typically found in hospital neuro-intensive care units, featuring an Intel® Core™ i3-1125G4 CPU (2.00 GHz, 4 cores, 8 logical processors) with a maximum clock speed of 2.0 GHz. This dual-system approach allowed us to assess computational feasibility in both research-grade and clinical bedside environments.

All computations were performed using Python (NumPy, Pandas, psutil).

#### • Statistics

To assess the effects of the different time windows on our metrics, we performed linear mixed-effects modeling (LMM) using the lme4 and lmerTest packages in R. The dependent variables included all the previously described metrics. The primary independent variable was the time window (window label), which categorized data into predefined phases (baseline, rise, and decay periods).

Each metric was modeled as a function of time window and a set of covariates related to sedative and vasoactive drug administration (Norepinephrine, Midazolam, Propofol, Ketamine, Thiopental, Esketamine, Remifentanil, Sufentanil, and Morphine). To account for inter-patient variability and repeated measures within individuals, we included a random intercept for each patient-event (n_event_patient). Given the hierarchical nature of our data—where each patient could experience multiple events, and each event was assessed across several time windows—LMM was preferred over traditional inferential methods such as ANOVA. Classical approaches assume independent observations and do not adequately account for within-subject correlations, which can lead to biased estimates in repeated-measures contexts. By including a random intercept, LMM allowed us to model subject-specific variability, capture intra-individual correlations, and improve the robustness of our statistical inferences. Additionally, LMM provides greater flexibility in handling unbalanced data, which is common in clinical studies where the number of observations per patient and time window can vary.

The general model structure was:

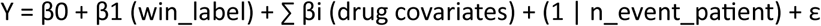

where Y represents the dependent variable, β0 is the intercept, β1 captures the fixed effect of the time window, drug effects are modeled as additional fixed effects, and a random intercept is included for each subject-event.

Model estimates were extracted, including β-coefficients, 95% confidence intervals (CIs), and p-values. Significance levels were categorized as p ≤ 0.001 (***), p ≤ 0.01 (**), p ≤ 0.05 (*), and non-significant (ns). The reference category for comparisons was Baseline Before (β = 0), with medians reported for descriptive purposes. Statistical analyses were performed in R (version 4.4.2), and results were compiled into structured tables for further interpretation.

#### ○ Spearman correlations in Python

To explore the relationships between time-domain and frequency-domain metrics derived from both intracranial pressure (ICP) and arterial blood pressure (ABP) signals, we computed Spearman’s rank correlation coefficient (R) using *scipy. stats. spearmanr()* function from SciPy. This non-parametric measure was chosen to assess potential associations between the different metrics. We applied this method to examine how variations in the time-domain and frequency-domain characteristics of ICP and ABP signals might be interconnected. To account for multiple comparisons, we applied a Bonferroni correction to adjust the p-values, reducing the risk of Type I errors.

## 3. Results

### • Dataset description

Study included 66 patients from 2020 to 2024 (40 males, 26 females) aged of 49.16 [38.38, 57.58] (median [1^st^ quartile, 3^rd^ quartile]) years old. They lasted for 7.03 [3.91, 11.76] days in the ICU where they were hospitalized mainly for SAH (51.52%), or TBI (33.33%), or other reasons (13.64). Their initial GCS was of 3.0 [3.0, 3.0] while their final was of 13.0 [9.0, 14.0], with an mRS score of 5.0 [4.0, 5.0].

Considering the 66 patients, ICP recordings lasted for 5.66 [3.22, 9.76] days, CO2 recordings lasted for 5.55 [3.45, 8.27] days, ABP recordings lasted for 5.6 [3.63, 9.09] days, ECG recordings lasted for 6.23 [3.68, 9.81] days.

### • Time-course of metrics during spontaneous ICP rise events

An automated pipeline allowed for the detection of 518 slow spontaneous ICP rise events. At least one event was detected in 56 of the 66 initially included patients. The number of events detected per patient was 6.0 [3.0, 12.0]. The total duration of these events was 118.45 [95.68, 164.97] minutes. This period was decomposed in a rising phase of 61.46 [44.82, 90.44] minutes that increased the ICP of 11.97 [9.01, 17.25] mmHg from 13.22 [9.57, 16.08] to 25.05 [22.23, 28.46] mmHg, followed by a decaying phase of 53.42 [40.55, 74.13] minutes that decreased the ICP of 10.88 [6.26, 16.05] to a value of 14.58 [10.45, 18.11] mmHg.

For analysis, rising and decaying phases were divided into 5 equal duration windows (+1 adjacent baseline window) of 12.0 [9.0, 18.0] and 11.0 [8.0, 15.0] minutes, respectively. Corresponding results are presented in Figure 2 and LMM results are available in supplementary table S1 with estimators and p-values.

**Figure 2:**
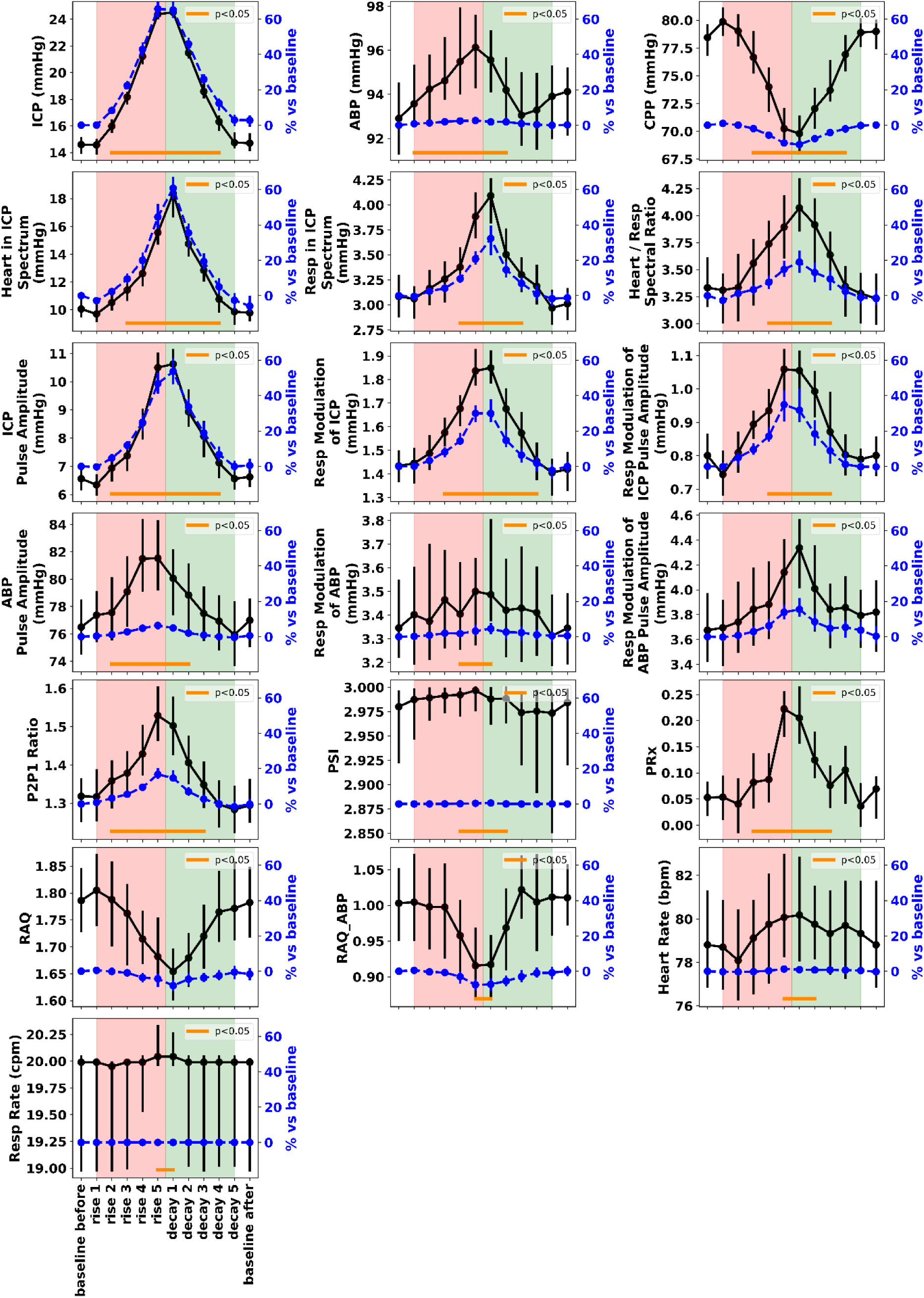
Point plots displaying the variation of metrics across time windows. Each subplot represents a different metric (name on the left), with central tendency estimated using the median for each time window. Error bars indicate 95% confidence intervals, computed via nonparametric bootstrap resampling. The black line represents data plotted in absolute units, while the blue dotted line represents data in relative units compared to the baseline (100 * (window_value – baseline_before_value) / baseline_before_value)). The orange horizontal line indicates where the estimators of the windows are statistically different (p < 0.05) from the baseline (baseline before), as assessed by linear mixed models.

As a control, we measured the median ICP level by window, which significantly increased, as described in the previous paragraph. Concurrently, the median ABP increased only slightly (from 92.91 [85.15, 101.65] to 96.13 [86.78, 106.07] mmHg, representing a change of +2.63 [-2.68, 8.64] %), likely in an attempt to maintain CPP. However, CPP decreased from 78.45 [71.13, 87.79] to 69.8 [61.23, 79.78] mmHg (-10.94 [-19.87, -2.44] %). The influence of the heart on ICP increased by 53.68 [24.07, 98.75] %, from 6.56 [4.62, 9.09] to 10.62 [7.19, 14.95] mmHg (ICP pulse amplitude), a finding almost identically observed in the frequency domain (Heart in ICP spectrum). The influence of respiration on ICP also increased, though to a lesser extent, by 30.02 [5.23, 76.87] %, from 1.43 [1.07, 1.83] to 1.85 [1.38, 2.5] mmHg (Respiratory Modulation of ICP), with almost identical relative changes observed in the frequency domain (Respiration in ICP spectrum). Consequently, the Heart/Respiratory spectral ratio (the ratio of the heart’s impact to the respiration’s impact in the frequency domain) also increased, though to a lesser extent (+19.02 [-21.05, 58.81] %). The respiratory modulation of ICP pulse amplitude also increased significantly, from 0.8 [0.56, 1.12] to 1.06 [0.68, 1.49] mmHg (+34.97 [-1.06, 77.33] %). We explored the dynamics of the same metrics in ABP, as it is closely linked to ICP. Both ABP pulse amplitude and the respiratory modulation of ABP increased, though only slightly (from 76.5 [64.06, 92.06] to 81.53 [68.45, 99.42] mmHg; +6.34 [-1.8, 16.03] % and from 3.35 [2.62, 4.64] to 3.5 [2.7, 4.86] mmHg; +3.14 [-9.26, 23.71] %, respectively), in the same direction as observed in ICP. Interestingly, respiratory modulation of ABP pulse amplitude increased slightly more than ABP pulse amplitude (from 3.67 [2.38, 5.57] to 4.14 [2.93, 6.45] mmHg; +13.8 [-5.6, 42.93] %), though not significantly. Probably due to the concurrent increases in both ICP and ABP, PRx rose from 0.05 [-0.15, 0.25] to 0.22 [-0.06, 0.47].The P2/P1 ratio increased from 1.32 [0.97, 1.66] to 1.53 [1.15, 1.94] (+16.72 [4.13, 32.87] %), while PSI marginally increased from 2.98 [2.24, 3.0] to 3.0 [2.46, 3.02] (+0.38 [-1.66, 16.96] %). Thus, PSI did not show significant variation, a point that will be discussed further in the Discussion section. RAQ2 showed a slight relative decrease (-8.13 [-25.74, 11.73] %), attributed to a smaller increase in the respiratory modulation of ICP (A_rp_ in the numerator) and a decrease in the respiratory modulation of ICP pulse amplitude (AA_vp_ in the denominator). A similar trend was observed for RAQ in ABP. Both heart and respiratory rates increased, but only marginally (by less than 1.25%).

### • Correlations of metrics during spontaneous ICP rise events

Thus, several metrics appear to covary concurrently during the detected challenges of ICP. We further explored these covariations using Spearman correlations and joint distributions of values. A subset of these relationships is shown in Figure 3. For the complete correlation matrix with associated statistical significance, refer to Supplementary Table S2, and for the full pairplot matrix with statistical significance, see Supplementary Figure S1.

**Figure 3:**
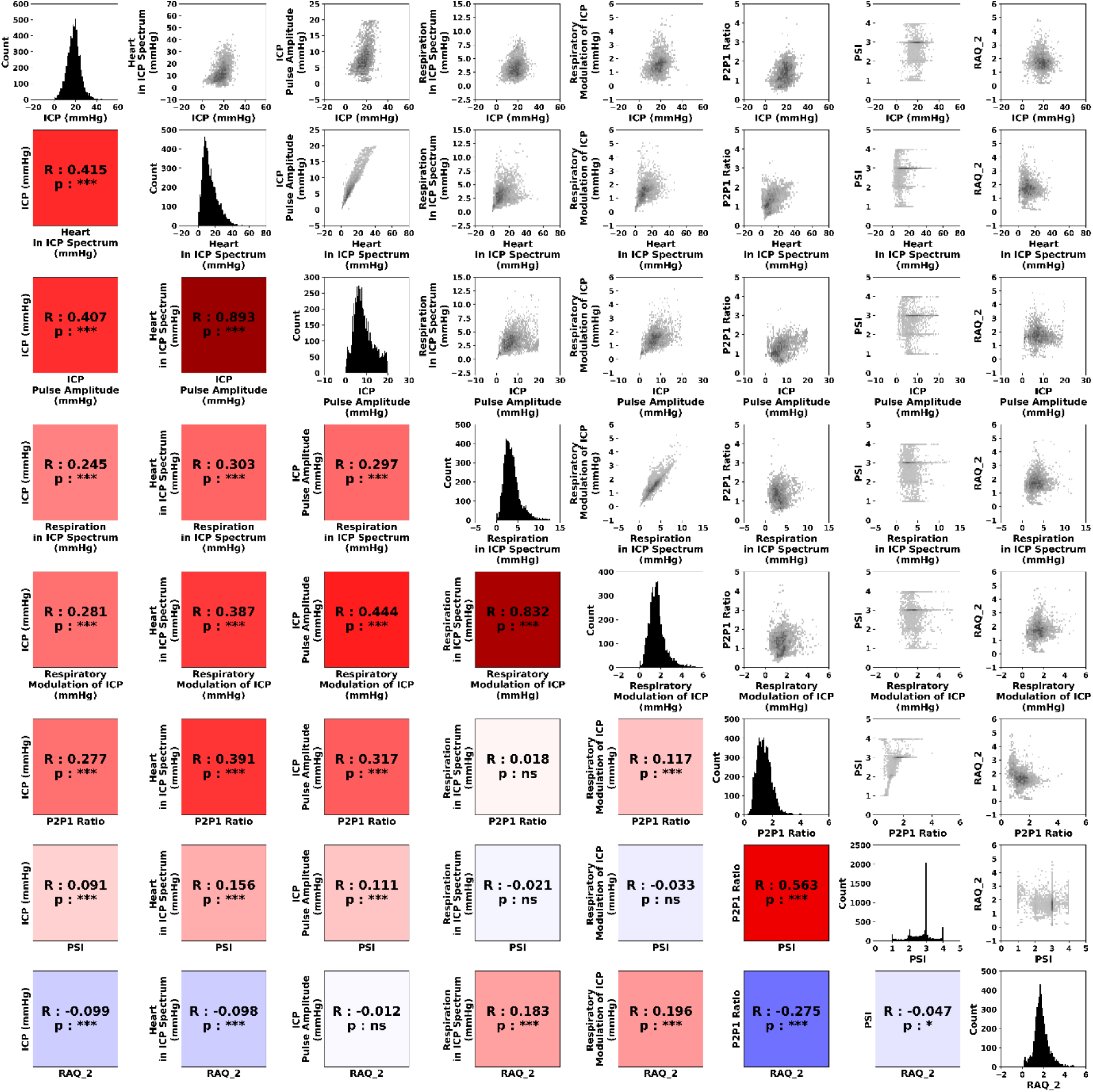
Pairplot and Correlation Matrix of Main Metrics. The lower-left triangle displays a correlation matrix using the Spearman method, where the R coefficient determines the color intensity, ranging from blue (-1) to red (1). Significance levels are indicated as follows: ns (p > 0.05), * (0.01 < p < 0.05), ** (0.001 < p < 0.01), and *** (p < 0.001). The upper-right triangle shows histograms representing the joint distribution of values. The diagonal displays the individual distribution of each metric.

Among the numerous correlations in the paired matrix, several are worth highlighting. First, as expected, the impact of the heart and respiration on ICP, assessed in the time domain, are strongly correlated with the same measures in the frequency domain (r > 0.8). The ICP level itself positively correlates with most of the metrics (except for RAQ2 and PSI), showing the strongest correlation with the impact of the heart on ICP (r = 0.415), though this is considered a moderate correlation. The P2/P1 ratio and PSI form a cluster of strongly correlated metrics (r = 0.563). Notably, the P2/P1 ratio moderately correlates with ‘Heart in ICP spectrum’ (r = 0.391), likely due to the fact that the assessment of ICP pulse amplitude, whether in the time or frequency domain, is based on the highest point of the pulse peak. In pathological situations, this peak may shift from P1 to P2, such that as P2 increases, while P1 remains almost fixed, both the P2/P1 ratio and the assessment of the heart’s measured impact on ICP also increase. Additionally, RAQ2 moderately anti-correlates with the P2/P1 ratio (r = -0.273).

### • Computational requirements of metrics

Results concerning the computational efficiency of the metrics are presented in Tableau 1. The process was iterated over one hour of data from 71 patients on two machines: one high-power and one low-power. Only the maximal values were retained, as these could overwhelm the computer and potentially cause crashes. It is worth noting that frequency domain metrics had the lowest computational cost, with one iteration taking less than 0.21 seconds, occupying less than 83% of one CPU, and consuming less than 10 megabytes of virtual memory. This allowed for the concurrent exploration of both heart and respiratory impacts on ICP. The slowest metric to compute was the P2/P1 ratio, taking less than 17 seconds per iteration. PSI, on the other hand, required the most computational resources, using over 4000% of CPU (equivalent to 40 cores at 100%) and 3.9 gigabytes of memory.

**Tableau 1:**
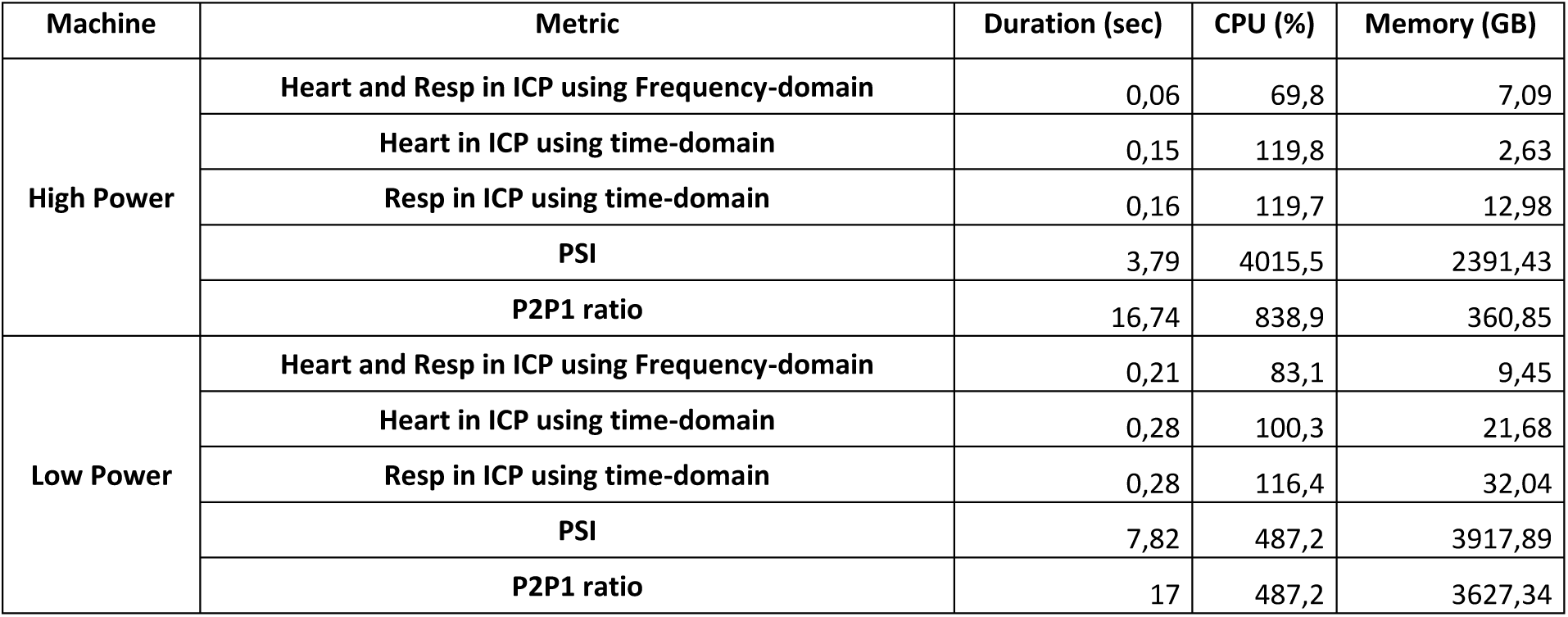
Computation requirements. Only the maximal values obtained from the computations iterated over the whole dataset were kept.

| Machine | Metric | Duration (sec) | CPU (%) | Memory (GB) |
| --- | --- | --- | --- | --- |
| High Power | Heart and Resp in ICP using Frequency-domain | 0,06 | 69,8 | 7,09 |
|  | Heart in ICP using time-domain | 0,15 | 119,8 | 2,63 |
|  | Resp in ICP using time-domain | 0,16 | 119,7 | 12,98 |
|  | PSI | 3,79 | 4015,5 | 2391,43 |
|  | P2P1 ratio | 16,74 | 838,9 | 360,85 |
| Low Power | Heart and Resp in ICP using Frequency-domain | 0,21 | 83,1 | 9,45 |
|  | Heart in ICP using time-domain | 0,28 | 100,3 | 21,68 |
|  | Resp in ICP using time-domain | 0,28 | 116,4 | 32,04 |
|  | PSI | 7,82 | 487,2 | 3917,89 |
|  | P2P1 ratio | 17 | 487,2 | 3627,34 |

## 4. Discussion

Our automated pipeline for detecting spontaneous slow ICP rises successfully identified 518 events in 56 patients, which likely challenged brain compliance. During these events, we examined several metrics potentially related to compliance by assessing the impact of heart and respiration on ICP in both the time and frequency domains. We found that the frequency domain metrics strongly correlated with time domain metrics during these challenges. In addition, the frequency domain metrics were much less computationally demanding. Finally, the AI-based analyses P2/P1 ratio and Pulse Shape Index formed a cluster of strongly correlated metrics (r = 0.563) that moderately correlated with the impact of the heart on ICP (r = 0.391 between the P2/P1 ratio and the heart component in the ICP spectrum).

### What did we detect?

The data from the ICCA software enabled us to get the date and approximate time of nurse interventions. We found that 45% of the detected events were influenced by at least one mobilization. A visual inspection of the raw ICP signal confirmed the sharp impact of such mobilizations on the ICP dynamics, which consequently increased. The remaining 55% of the detected ICP rises were likely caused by spontaneous awakenings or plateau waves of ICP. (Dias et al., 2016; Kasprowicz et al., 2016; Risberg et al., 1969).

### What are the most sensitive metrics to our brain ICP challenge events?

From a global perspective, our analysis indicates that most of the computed metrics are positively correlated with the ICP level itself, except for the PSI and RAQ2. Among all the metrics, the impact of the heart on ICP (i.e., ICP pulse amplitude) is the most sensitive to increases in ICP (r > 0.4), while the impact of respiration on ICP also shows a positive correlation, albeit to a lesser extent (r > 0.4).

Interestingly, this is also true for the P2/P1 ratio. This could likely be due to the fact that the assessment of ICP pulse amplitude (i.e., the impact of the heart on ICP), whether in the time or frequency domain, is based on the highest point of the pulse peak. In pathological conditions and/or in cases of high ICP, this peak may shift from P1 to P2. As a result, while P1 remains almost fixed, an increase in P2 leads to both an increase in the P2/P1 ratio and in the assessment of the heart’s impact on ICP.

RAQ2, our potentially more accurate version of RAQ, showed almost no correlation with the ICP level. This is not surprising, as it was specifically designed to reflect alterations in the pressure-volume relationship without being influenced by changes in ICP (Spiegelberg et al., 2020).

PSI showed almost no correlation with the ICP level. Furthermore, PSI correlated poorly with any metric, except for the P2/P1 ratio (r = 0.563), and to a much lesser extent with the impact of the heart on ICP (r = 0.156). The correlation between PSI and the P2/P1 ratio is expected, as both metrics aim to extract information from the ICP waveform induced by the cardiac pulse. This waveform primarily depends on the relative positions of P1, P2, and P3, which determine the classification of the pulse type into 1,2,3 or 4^th^ pulse type (Kazimierska et al., 2023; Mataczyński et al., 2022). One might wonder why these metrics are not more strongly correlated. First, noise in the ICP signal could affect the computation of the P2/P1 ratio or the classification of the pulse shape, even though these are encoded to reject or smooth out noisy detections. Additionally, pulse shape classification could benefit from other information beyond the P1 and P2 peaks for more accurate classification. However, the main limitation may lie in the PSI scoring itself, specifically the lack of continuity in its distribution. Seeing its distribution in Figure 3 highlights this statement. We observe that its values tend to cluster around integers, which is expected given that the model classifies using integer values. However, one might question the relevance of this statistical tendency to ‘integer’ the values, considering that a theoretically exact measure of brain compliance should be a true continuous variable. Therefore, PSI may serve as a useful tool to guide a clinician’s judgment, though it may not be as precise as one might expect.

Finally, can the volumetric changes imposed by respiration and cardiac activity on the intracranial space be used to assess brain compliance? The intracranial volume is traditionally considered the sum of parenchymal volume, blood volume, cerebrospinal fluid (CSF) volume, and, in pathological conditions, mass lesion volume. In healthy humans, the respiratory cycle accounts for approximately 0.8 mL of cerebral blood volume variation, while the cardiac cycle contributes about 1 mL (Söderström et al., 2025). CSF flow is largely driven by both cardiac and respiratory rhythms, with peak volumetric flow rates in the cerebral aqueduct recorded in average between 0.3 and 0.4 mL/s for both respiratory and cardiac components (Vinje et al., 2019). The brain parenchyma itself may also undergo volumetric changes in response to respiratory and cardiac activity, given that neuronal activity—and likely glial cell activity—has been increasingly described as being partially influenced by these physiological cycles (Heck et al., 2017; Juventin et al., 2022; Park & Blanke, 2019). Intracranial volume is therefore modulated by cardiac and respiratory cycles, though with significant intra- and inter-individual variability (Söderström et al., 2025). Such natural volume variations should be buffered in conditions of high brain compliance but less so in cases of low compliance. In our study, we showed that as ICP increased, the impact of both cardiac and respiratory components on ICP also increased (r > 0.4), presumably due to a decrease in brain compliance and a shift to the right of the pressure volume curve as suggested by the decrease in the RAQ. However, further investigations combining measurements of heart- and respiration-induced intracranial volume variations with corresponding intracranial pressure variations could provide deeper insights into this phenomenon, as has been partially explored in previous studies (Söderström et al., 2025; Vinje et al., 2019).

### About the practical usability of metrics

Our analysis of computational requirements revealed that frequency-domain assessment of the cardiac and respiratory impact on ICP was by far the most computationally efficient approach. In contrast, machine learning-derived metrics (such as the P2/P1 ratio and PSI) were the least efficient, even when using pre-trained models. Regarding the latter, and considering that our tests were conducted on only a one-hour sample signal iterated over the entire dataset, we observed reasonable computation times (on the order of tens of seconds) but with memory consumption reaching up to 4 GB. This is an important consideration for deployment in the intensive care unit (ICU) environments, where computational resources are often limited. Further research should explore the relationship between the continuous intracranial volume fluctuations induced by cardiac and respiratory cycles and brain compliance. If a strong and clinically relevant link is established, then frequency-domain analysis would likely be the optimal strategy, offering a fast and lightweight means of assessing brain compliance-related information. Lastly, we believe it is worth noting that methods using ICP waveforms were developed based on intraparenchymal pressure sensors, rather than intraventricular ones, the latter being the gold standard for intracranial pressure monitoring (R. Chesnut et al., 2014), whereas the waveform can drastically change between the two, possibly leading to miscalculations of PSI or the P2/P1 ratio. The frequency-domain Fourier method, which involves sinusoidal convolutions, could be less biased by such waveform variations and therefore provide more reproducible results depending on the sensor placement area. Additional research should be conducted to confirm this point.

### Relationships with ABP

Our results showed that during the detected ICP rises, ABP increased slightly. Consequently, PRx also increased, which could be interpreted as a disruption of cerebral flow autoregulation. However, it should instead reflect a proper adaptation (spontaneous, or secondary to medical intervention) of ABP to the ICP rise to maintain CPP, suggesting that PRx may not be reliably interpretable when changes in CPP result from variations in ICP rather than ABP. Thus, we argue that PPCopt deduction (from PRx and PPC) is not interpretable in situations where CPP decreases are driven by ICP increases rather than ABP decreases—a frequent occurrence, as observed in our study.

We then explored the dynamics of the same metrics developed for ICP but applied to ABP. ABP pulse amplitude, respiratory modulation of ABP, and respiratory modulation of ABP pulse amplitude increased slightly as ICP rose. Interestingly, among these three metrics, respiratory modulation of ABP pulse amplitude exhibited the greatest relative increase. Furthermore, what we termed RAQ_ABP—analogous to the RAQ developed for ICP but applied to ABP—was approximately equal to 1, whereas for ICP, it was approximately 1.8. This indicates that in ABP, the A_rp_ component (respiratory modulation of ABP) is nearly equivalent to the AA_vp_ component (respiratory modulation of ABP pulse amplitude). Specifically, respiratory modulation of ABP was 2.2 times that of ICP, while respiratory modulation of ABP pulse amplitude was 4 times that of ICP pulse amplitude. Therefore, it appears that respiratory modulation of pulse amplitude is not transferred from ABP to ICP dynamics as effectively as the respiratory modulation of the signal level itself. Consequently, the documented association between respiratory modulation of ABP pulse amplitude and preload dependency (Bronzwaer et al., 2015) may represent a hemodynamic state that is buffered within the intracranial compartment. This interpretation is supported by the small yet significant correlation between ICP and ABP pulse amplitude (r = 0.224, p < 0.001; see Supplementary Figure S1 and Table S2) while, in contrast, respiratory modulation of ICP and ABP levels exhibited a much stronger correlation (r = 0.637, p < 0.001).

### How can the clinician use such metrics

Considering these results, we believe that potential changes in cerebral blood volume, as assessed by our metrics in response to different CPP levels, are particularly relevant when cerebral compliance is abnormal. The ICP waveform, P2/P1 ratio, and ICP pulse amplitude could serve as surrogate markers of cerebral compliance. We argue that these metrics should be evaluated before testing cerebral autoregulation. Indeed, if cerebral compliance is normal (i.e., P2 < P1, low PSI, and low ICP pulse amplitude), changes in CPP do not induce significant cerebral blood volume changes, rendering PRx uninformative. Conversely, an increase in any of these metrics—even if ICP remains within the normal range—should raise concern and indicate the need for cerebral autoregulation assessment. Furthermore, these metrics may be valuable during the step-down phase of ICU treatment, such as sedation withdrawal. Their “normality” could suggest a safe opportunity for withdrawal attempts, whereas abnormalities might indicate the need to postpone such interventions.

Finally, we propose an overview helper tool in Figure 4, that summarizes key features relevant for clinicians to monitor a patient’s cerebral condition, at least based on ICP. Notably, the ICP waveform view alone (left panels) offers sufficient insight into the patient’s brain status, encapsulating the metrics shown in the right panels. Indeed, a simple visual inspection of the average ICP waveform provides valuable information on 1) the quality of the ICP signal, 2) the ICP level, and 3) pulse shape characteristics (P2/P1 ratio, PSI, and pulse amplitude).

**Figure 4:**
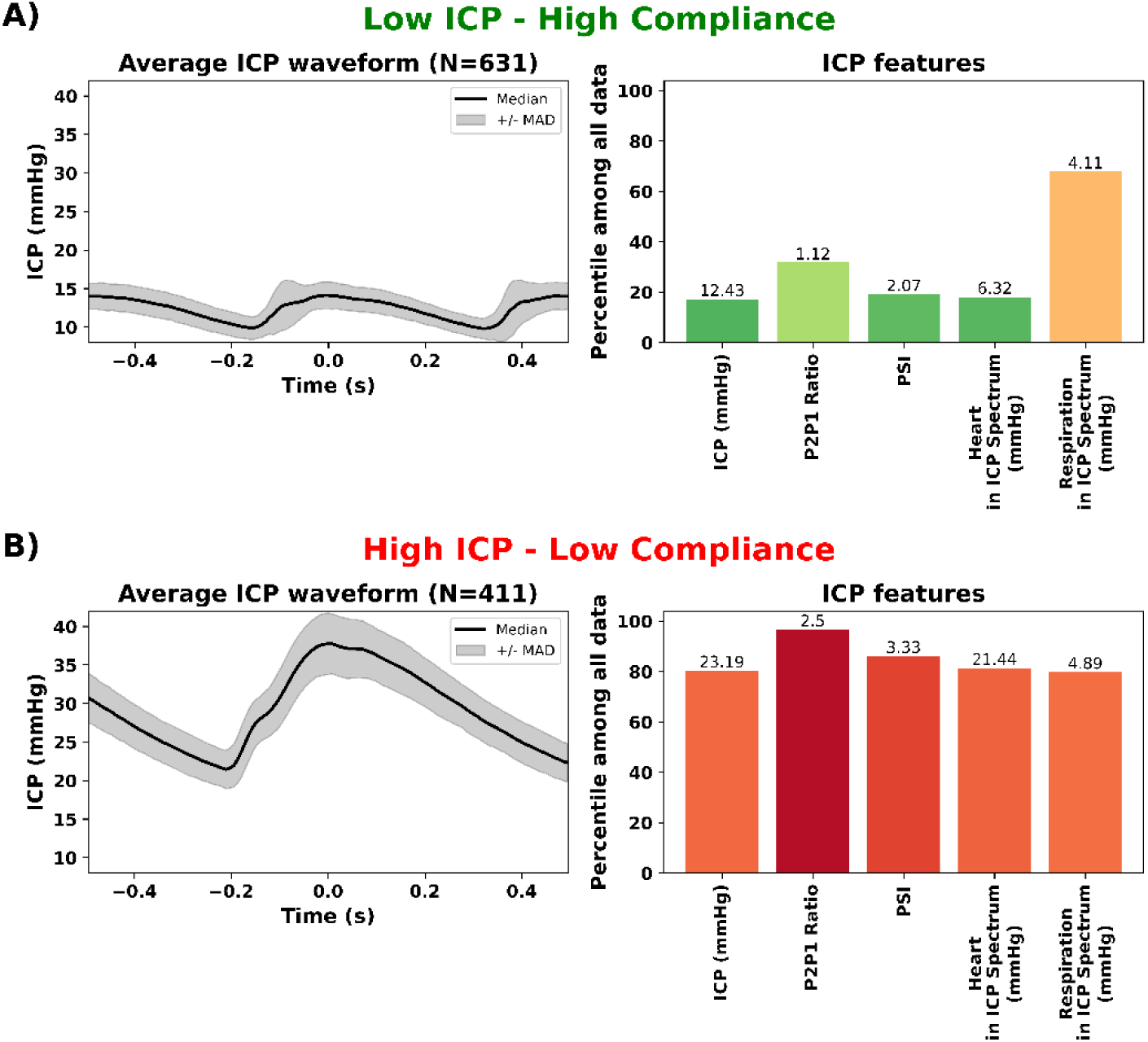
ICP Overview Helper Tool. A) Example of an ICP overview helper for clinicians, showing features derived from 5 minutes of ICP signal sampled during a low ICP and high compliance period. The left panel displays the average waveform (median of the epochs) with a surrounding shading representing statistical dispersion (± median absolute deviation – MAD). The right panel is a bar plot illustrating percentiles of current values for clinically relevant features (ICP level, P2P1 ratio, Pulse Shape Index, and the impact of heart rate and respiration on ICP), sorted among a large dataset. Bar colors range from green, indicating clinically normal values based on the metric rank, to red, indicating clinically concerning conditions. B) Same features derived from 5 minutes of ICP signal sampled during a high ICP/low compliance period. Note that the ICP waveform view alone (left panel) provides sufficient insights into the patient’s brain condition, summarizing the metrics displayed in the right panel.

## Conclusion

This study introduces an automated pipeline for detecting spontaneous slow ICP rises and explores several metrics related to brain compliance. While the impact of cardiac and respiratory activity on ICP provides valuable insights, it should be considered as supplementary to, perhaps, more direct indicators. ICP waveform metrics, such as the P2/P1 ratio and pulse amplitude, show strong correlations with ICP levels and likely brain compliance, offering practical utility for clinicians in assessing brain condition. Notably, frequency-domain analyses, which assess the impact of cardiac and respiratory activity on ICP, provide nearly the same informative value as more complex machine learning-based tools, but with the advantage of being much less computationally demanding. This makes them particularly suitable and intuitive for real-time clinical monitoring in hospital settings, where computational resources are often limited. Overall, ICP waveform analysis—combining both spectral analysis and advanced techniques—offers a robust and efficient approach for evaluating brain compliance in clinical practice.

## Acknowledgments

We would like to express our sincere gratitude to the caregivers at HCL (Hospices Civils de Lyon) for their support. We also thank the computer scientists at CRNL (Centre de Recherche en Neurosciences de Lyon) for developing the valuable computing environment that enabled us to process large data sets. Special thanks to Donatien Legé for the insightful discussions that shaped this work. Finally, we acknowledge the staff at Moberg for developing the hardware and software that were essential for recording the data used in this study.

## Data availability

The datasets reported herein are available from the corresponding author on a reasonable request.

## Code availability

The code used in this study includes contributions from multiple sources:

- **physio Toolbox**: Available at https://github.com/samuelgarcia/physio
- **pycns Toolbox**: Available at https://github.com/samuelgarcia/pycns
- **Project-Specific Code**: https://github.com/ValentinGhibaudo/Scripts_Compliance_Project

## Abbreviations

ICU: Intensive Care Unit
ICP: Intracranial Pressure
GCS: Glasgow Coma Scale
mRS: Modified Rankin Scale
DC: Direct Current
Hz: Hertz
PSI: Pulse Shape Index
CPU: Central Processing Unit
SAH: Subarachnoid Hemorrhage
TBI: Traumatic Brain Injury
ABP: Arterial Blood Pressure
ECG: Electrocardiogram

## Supplementary

**Figure S1.**
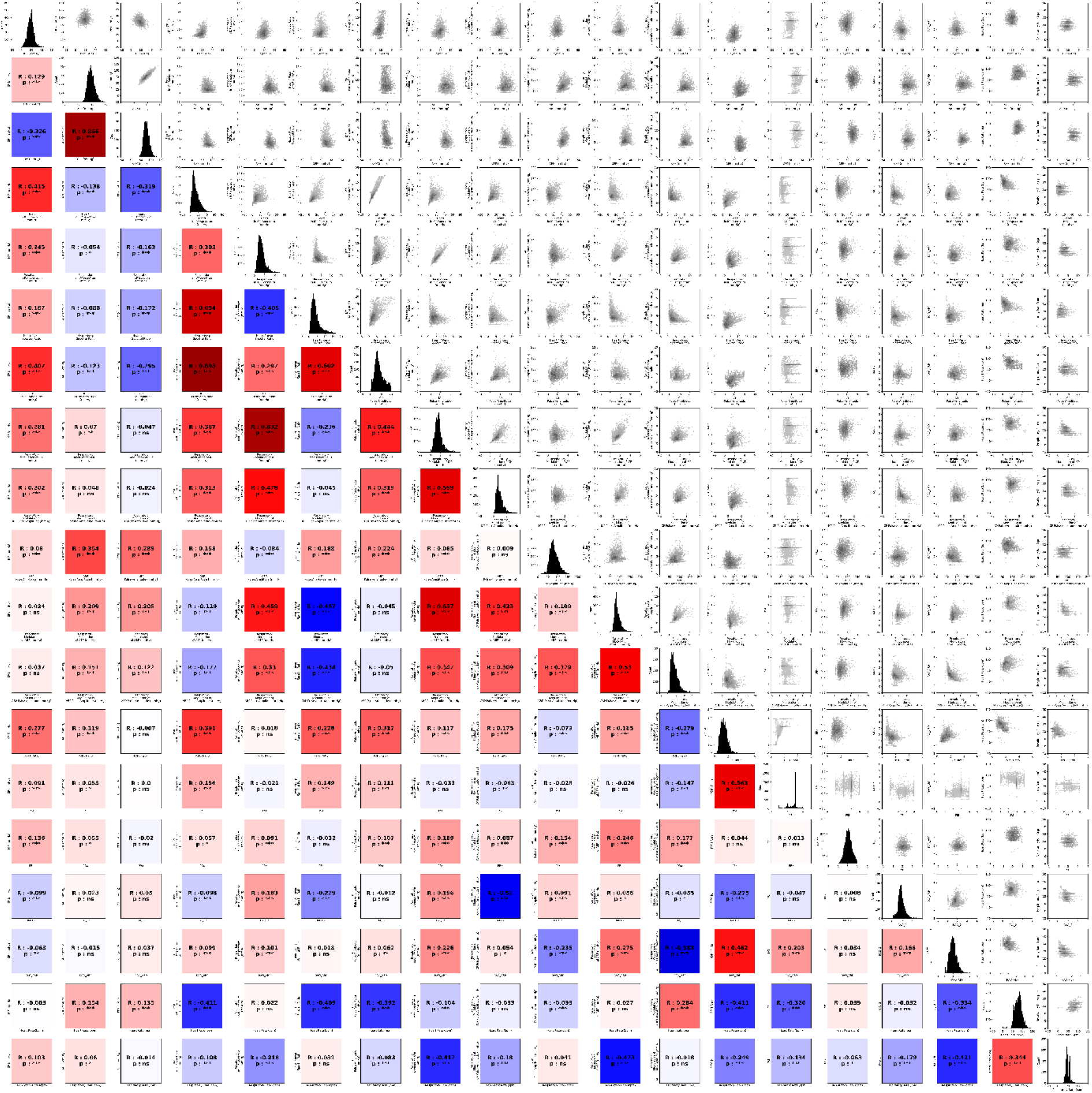

**Table S1.**
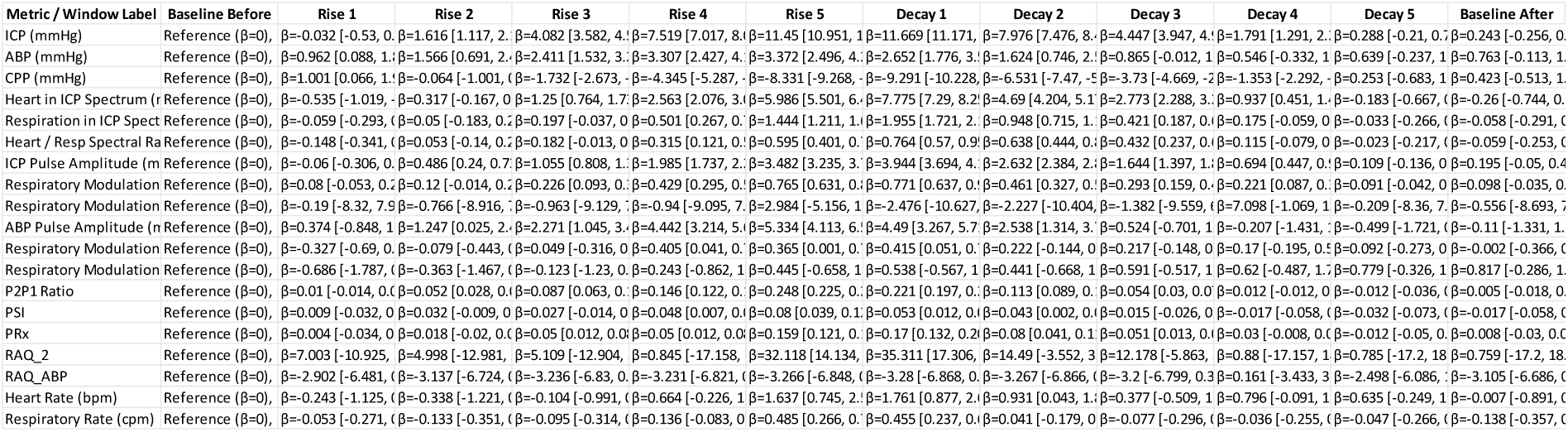

**Table S2.**
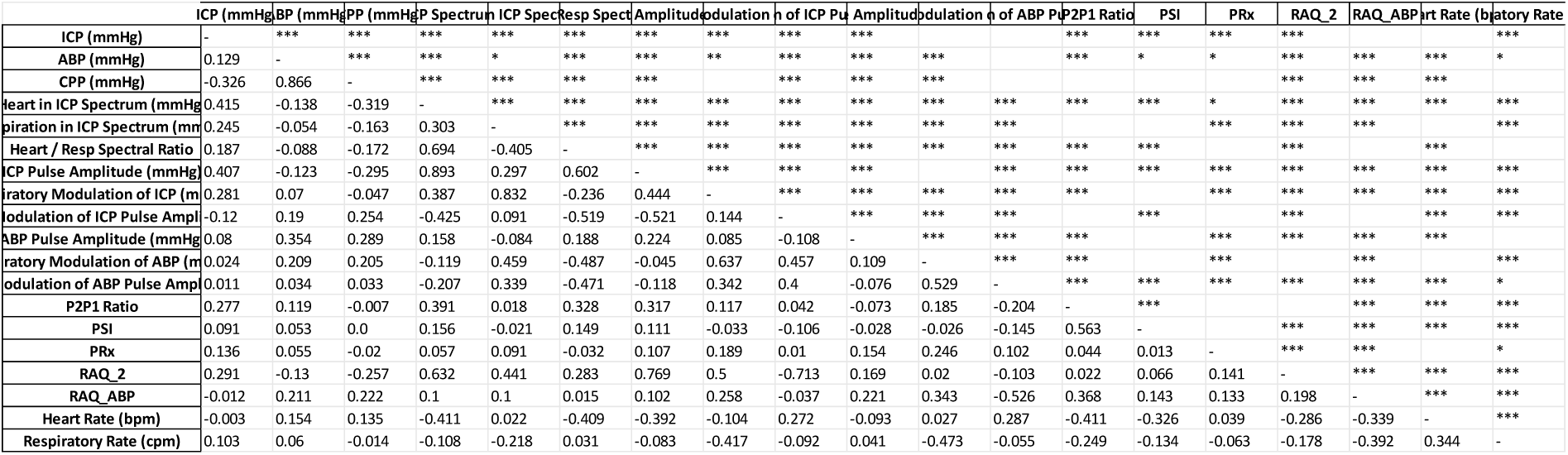

